# iMDPath: Interpretable Multi-task Digital Pathology Model for Clinical Pathological Image Prediction and Interpretation

**DOI:** 10.1101/2025.04.13.25323912

**Authors:** Qitao Chen, Zhe Wang, Xia Lin, Yuying Shi, Botao Xu, Jie Chai, Tao Zhang, Cheng Wang

## Abstract

Deep learning (DL)-based pathological image modelling and analysis approaches offer transformative potential for early cancer diagnostics, yet limited sample sizes and a lack of interpretability often hinder efficient clinical translation. Here, we present the interpretable Multi-Task Digital Pathology Model (iMDPath), an end-to-end highly explainable multi-task deep learning framework that simultaneously addresses these challenges by integrating data augmentation, diagnostic prediction, and visualization of pathological image features. The iMDPath comprises three modules: Augmentation (iMDPath-Aug), Prediction (iMDPath-Pred), and Visualization (iMDPath-Vis). iMDPath-Aug incorporates a vector-quantized variational autoencoder (VQ-VAE) for enhanced data augmentation, capturing essential pathological features from limited datasets. A Swin Transformer-Based (Swin-B) predictor in the iMDPath-Pred module leverages the augmented data to achieve better performance than state-of-the-art models across four diverse cancer pathology datasets, including gastric and breast cancer. Finally, iMDPath-Vis, a novel visualization module combining the full gradient (FullGrad) and occlusion sensitivity analysis, provides pathologists with actionable insights by highlighting the specific tissue regions driving model predictions. Overall, iMDPath not only surpasses existing methods in diagnostic accuracy, sensitivity, and generalization across these datasets, but also offers a transparent and interpretable AI solution for precision oncology, paving the way for more reliable and efficient clinical decision-making.

## Introduction

Pathological images are crucial for clinical diagnostics in cancer, providing essential information for cancer grading and progression^1-3^. Evaluating the cellular features based on the pathological images not only enhance the accuracy of cancer diagnosis but also helps inform therapeutic decisions, shaping personalized treatment strategies, which can significantly improve patient outcomes^4,5^. However, traditional manual microscopic evaluation faces challenges including inter-observer variability and time-intensive analysis. Given these limitations, Deep learning (DL)-based approaches that can effectively utilize pathological images hold significant promise to advance diagnostic accuracy and clinical workflows. Over the past decade, DL-based approaches have been increasingly developed to facilitate the analysis of pathological images, enabling rapid processing of large-scale datasets, identifying microscopic lesions, and achieving high precision in disease classification^6-8^. In addition, large-scale foundation models have emerged to enhance clinical decision-making based on clinical pathological images, offering valuable support to pathologists and physician scientists in optimizing therapeutic strategies^9-11^.

Among these developments, several landmark studies have demonstrated the versatility of different DL architectures in pathological applications. Kumar et al.^12^ utilized the Visual Geometry Group (VGG) network for the Multi-Organ Nucleus Segmentation Challenge, demonstrating its ability to extract precise features from pathological images. Marme et al.^13^ applied the Residual Neural Network (ResNet) to predict breast cancer sentinel lymph node status, employing skip connections to enhance training depth and accuracy. Coudray et al.^14^ leveraged the Inception network for classifying and predicting mutations in non-small cell lung cancer by utilizing its multi-scale feature processing. Ghaffari et al.^15^ benchmarked Vision Transformer (ViT) for whole-slide classification, utilizing its global attention mechanism to address challenges associated with limited data. Despite these studies highlighting the potential and applicability of DL models in pathological image-based diagnosis, there remains a critical need for extensive, diverse training datasets that represent the full spectrum of lesion types encountered in clinical practice for developing accurate models^16-22^. Limited sample sizes in training the accurate model remain a challenge in developing the customizable and explainable pathological image deep learning models. In addition, the desirable DL models must be robust and generalizable to handle the variations in pathological imaging techniques and staining protocols across different institutions. The lack of interpretability hampers the ability to understand the relationship between the pathological images and disease progression^23^.

To address these limitations, we developed the interpretable Multi-Task Digital Pathology Model (iMDPath), an end-to-end framework designed to perform multiple tasks in pathological image modelling and analysis, including dataset augmentation, disease grade prediction, and interpretation of pathological image regions of interest. The iMDPath framework comprises three key modules: Augmentation (iMDPath-Aug), Prediction (iMDPath-Pred), and Visualization (iMDPath-Vis). The iMDPath-Aug module employs a variational autoencoder-based model for image augmentation, improving the representation of pathological features. The iMDPath-Pred module utilizes a Swin Transformer-Based (Swin-B) architecture to enhance prediction accuracy for a wide range of cancer pathological images. The iMDPath-Vis module provides interpretability by generating visualizations that highlight key regions of interest aligned with the model’s predictions. By applying iMDPath framework to four different pathological datasets, the iMDPath framework not only achieves excellent prediction performance compared to state-of-the-art models, but also offers instructional insights regarding the critical regions that contribute to the model’s predictions. The iMDPath exhibits great potential for scaling up the generalizability across larger and more diverse datasets to ensure its robustness and applicability in various clinical settings. The iMDPath provides a comprehensive tool for assisting pathologists in digital pathology, enhancing the clinical utility of DL models in precision oncology.

## Materials and Methods

### Datasets

In this study, we utilized four pathological datasets of gastric and breast cancer from multiple data sources, including BOT dataset, SEED dataset, SDGC dataset, and BreaKHis dataset. The BOT (Brain of Things, http://www.datadreams.org/) dataset was obtained from the 2017 China Big Data Artificial Intelligence Innovation and Entrepreneurship Competition, including 520 pathological images of gastric cancer classified into malignant and benign categories. The SEED dataset was collected from the Big Data Development and Application Competition (Medical and Health Track, https://www.jseedata.com), containing 743 gastric cancer images categorized into normal, tubular, and mucinous conditions. The SDGC dataset was collected from Shandong Cancer Hospital and contained 753 images of gastric cancer, including four categories: moderately differentiated (225 images), poorly differentiated (273 images), poorly differentiated with signet-ring cells (193 images), and signet-ring cells (62 images). The BreaKHis (Breast Cancer Histopathological Image Classification) dataset contained 7,909 small biopsy images from 82 patients diagnosed with breast cancer, categorized into benign and malignant tissues^24,25^.

### Interpretable Multi-Task Digital Pathology Model Framework

The iMDPath framework comprises three core modules: an augmentation module, a prediction module, and a visualization module. The iMDPath-Aug module enhances image quality using a variational autoencoder model. The iMDPath-Pred module, powered by the Swin-B architecture, improves the prediction accuracy of pathological images. The iMDPath-Vis module generates visualizations of the predicted outcomes by highlighting key tissue regions in the pathological images, providing interpretability for the model’s predictions.

### Augmentation for Pathological Images

The iMDPath-Aug module employs the vector-quantized variational autoencoder (VQ-VAE)-based augmentation to address the challenge of limited pathological image data^26^ (**Figure 1A**). This module incorporates a vector quantization mechanism to enhance the capture of critical pathological features. The iMDPath-Aug module consists of three components: an encoder, a vector quantization layer, and a decoder.

**Fig. 1.**
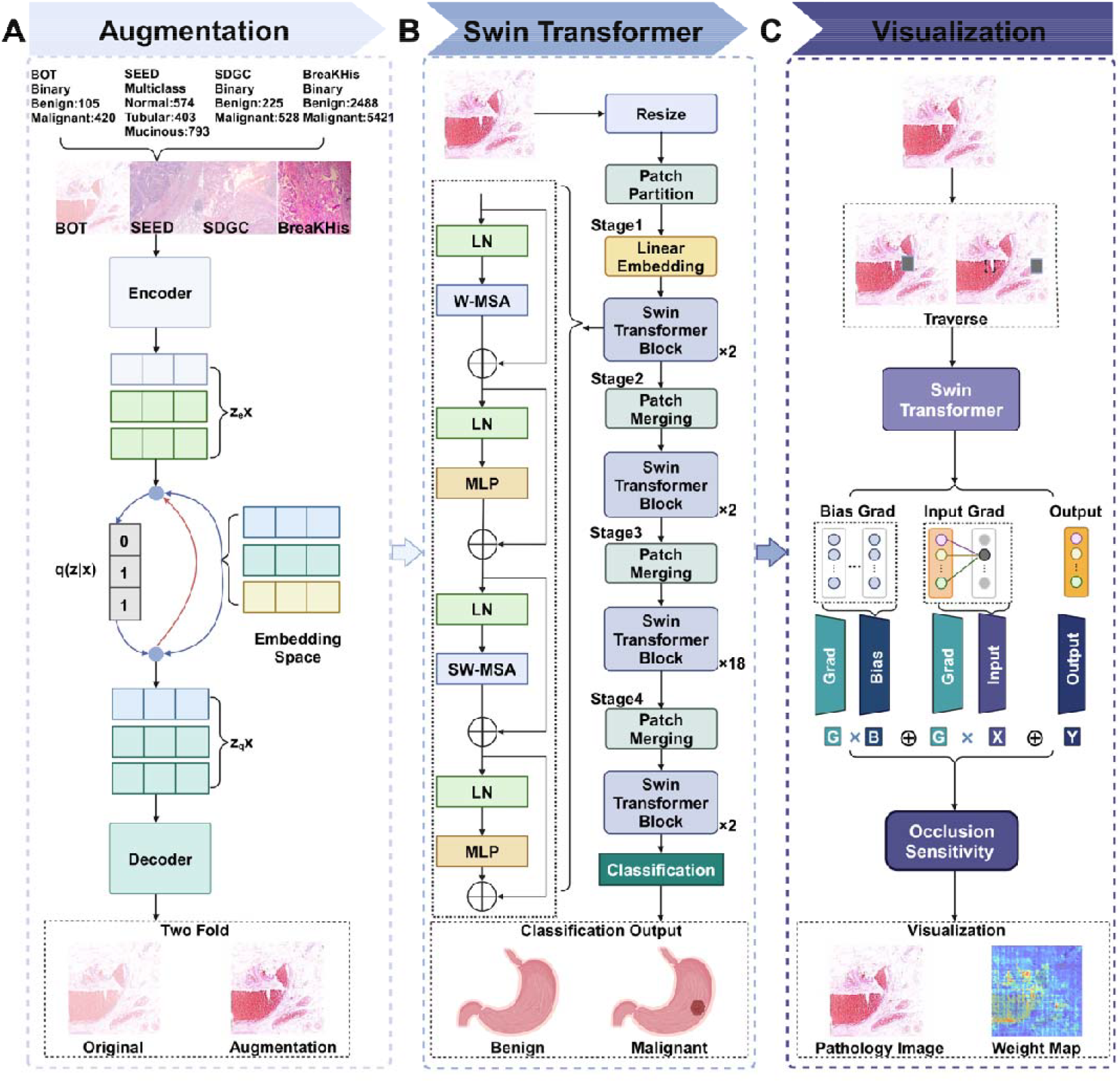
Framework of the interpretable Multi-Task Digital Pathology Model (iMDPath). Panel A illustrates the iMDPath-Aug model that implements VQVAE-based augmentation process, which enhances pathological images through an encoder, vector quantization layer, and decoder, improving image quality and consistency. Panel B depicts the iMDPath-Pred module, which optimizes Swin-B model that divides images into patches and captures multi-scale features for accurate outcome prediction across diverse datasets. Panel C demonstrates the explanation module iMDPath-Vis, combining the FullGrad and occlusion sensitivity to generate heatmaps that highlight key regions influencing prediction outcomes.

The encoder extracts features from 512×512 RGB pathology images, progressively downsampling them through convolutional layers to a latent feature space. The feature mapping can be represented as:

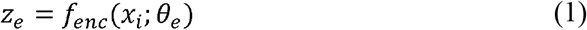

where *x*_*i*_ is the input pathology image, *z*_*e*_ is the learned *m* dimensional feature representation, and *θ*_*e*_ is the set of parameters of the encoder.

Subsequently, the vector quantization layer discretizes the encoded features *z*_*e*_ by mapping them to the closest vector in a predefined codebook *e*_*k*_:

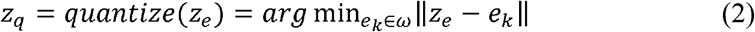

where *ω* is the set of codebook vectors, and *e*_*k*_ is a vector from the codebook.

The decoder then reconstructs the input pathology image 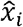 from the quantized latent representation *z*_*q*_:

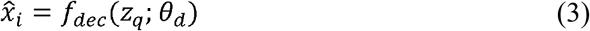

where *θ*_*d*_ represents the decoder’s parameters, and 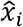 is the reconstructed image.

To ensure the generated images maintain critical pathological features while achieving robust enhancement, we employ the total loss function *L* that combines reconstruction loss, vector quantization loss, and commitment loss. It is defined as:

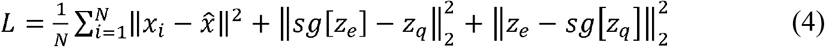

where *sg*[·] is the stop-gradient operator and *N* is the batch size.

Together, these efforts result in a flexible and high-performance tool for pathological image analysis, providing clinicians with more accurate, efficient, and interpretable diagnostics tailored to meet the critical needs of precision oncology.

### Generalization for Diagnosis Prediction

Pathological image analysis poses challenges due to the variability across different clinical institutions and the noise in the data. To address these, the iMDPath-Pred module leverages the Swin-B architecture (**Figure 1B**), which consists of four stages with 24 transformer blocks. The Swin-B uses shifted window attention to capture both local and global features, making it effective for identifying diagnostic patterns at multiple scales^27^.

The iMDPath-Pred module processes input pathological images represented as 4D tensors with dimensions (B, H, W, C), where B is the batch size, H and W are the spatial dimensions, and C=3 represents the RGB channels. Through an initial patch partition step, these images are divided into non-overlapping 4×4 patches^28^. These patches are projected into a higher-dimensional feature space through linear embedding, generating feature maps of shape (B, H/patch size, W/patch size, C). Next, feature maps undergo progressive merging through the four stages, with feature dimensions of 128, 256, 512, and 1024 respectively. Each swin transformer block includes layer normalization (LayerNorm), window-based multi-head self-attention (W-MSA), shifted window multi-head self-attention (SW-MSA), and a multi-layer perceptron (MLP). While W-MSA captures local features within defined windows, SW-MSA employs window shifting to enable global feature extraction, enhancing the model’s capability to process diverse pathological patterns. Residual connections improve training stability and mitigate vanishing gradients, producing rich feature maps essential for diagnosis. The subsequent patch merging further captures multi-scale features by concatenating adjacent patches, reducing spatial resolution and increasing feature dimensionality. A 1×1 convolution is applied to manage complexity while retaining critical diagnostic information.

The final feature map is globally pooled and passed through a softmax function for classification, effectively combining local and global information. We employed transfer learning to train the Swin-B model. Pre-trained weights from ImageNet were used to initialize the network, with the classification head fine-tuned on the dataset. The model was further optimized by unfreezing all layers and refining performance using the stochastic gradient descent (SGD) optimizer with a learning rate of 0.001. We trained the model for 100 epochs to minimize cross entropy loss, with the model achieving the lowest validation loss selected for evaluation. This approach ensured robust diagnostic performance tailored to pathological image analysis.

### Visualization of Latent Embeddings on Pathological Images

To enhance the transparency and interpretability of diagnostic results in clinical applications, we developed the iMDPath-Vis module by integrating the full gradient (FullGrad) and occlusion sensitivity analysis^29^ (**Figure 1C**). The FullGrad calculates pixel-level contributions by computing the gradient of the model’s output *f* (*x*) with respect to the input pathology image *x*. This gradient, 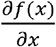, highlights how each pixel influences the prediction. The contribution of each pixel is emphasized by weighting the gradient with the input image values:

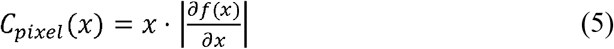

Additionally, gradients of the model’s bias terms *b*_*l*_ at each layer *l* are calculated as 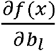 and weighted by the bias values to quantify their contribution:

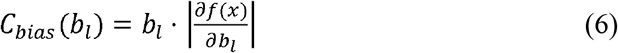

The FullGrad class activation map (CAM) aggregates these contributions across all layers and channels to identify key regions in the image:

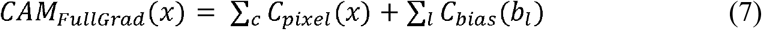

In parallel, occlusion sensitivity analysis systematically masks regions of the input image *x* using a sliding window of size 7×7 and stride 3 (aligned with the Swin-B). For a window centered at position, *v*, the occluded image *x*_*occlude*_ is defined as:

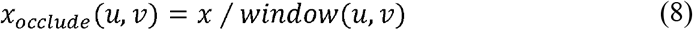

The sensitivity of the prediction is quantified by the change in classification score:

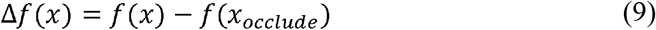

The occlusion-adjusted the FullGrad CAM is recalculated for each occluded image, *CAM*_*occlude*_ (*x*) . The final sensitivity map combines the score change and CAM change as a weighted sum:

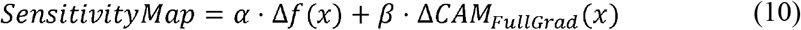

where *α* and *β* are weights controlling the relative contributions of score and CAM changes. This method generates interpretable heatmaps that highlight critical regions affecting the model’s prediction.

### Model Training and Performance Evaluation

During model training, all pathological images from the four datasets were preprocessed prior to input into the iMDPath framework. Each image was standardized and resized to 512×512 pixels for the iMDPath-Aug module and to 224×224 pixels for the other two modules. The datasets were then split into training (70%), validation (20%), and test (10%) sets to ensure proper model training and testing. To rigorously evaluate the model’s performance, multiple evaluation metrics were employed, including loss curves, accuracy, confusion matrices, area under the curve (AUC), receiver operating characteristics (ROC), precision-recall (PR) curves, and average precision (AP). The ROC curve was plotted by comparing the true positive rate (TPR) against the false positive rate (FPR). The PR curve was constructed by calculating and plotting precision and recall across various thresholds. The detailed evaluation functions are listed as follows:

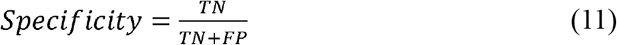

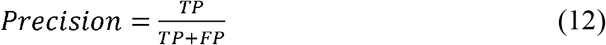

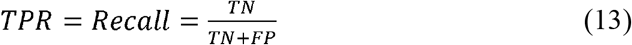

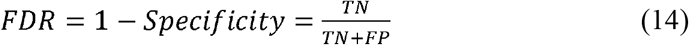

where TP represents true positive, FP represents false positive, TN represents true negative, and FN represents false negative.

### Comparison between iMDPath and Baseline Models

This study implemented several baseline models to systematically compare and evaluate the performance of the modules in the iMDPath framework. For the iMDPath-Aug module, we utilized traditional image preprocessing techniques and the variational autoencoder (VAE) as baseline approaches to assess the effectiveness of the proposed VQ-VAE for data augmentation performance^30^. The traditional preprocessing methods, including random cropping, resizing, horizontal flipping, color jittering, and rotation, served as key reference points, enabling the evaluation of the iMDPath-Aug module’s ability to enhance image diversity and robustness. Additionally, the VAE, as a control generative model, provided a comparative framework to assess the potential benefits of integrating vector quantization in the image enhancement process. For the iMDPath-Pred module, we selected VGG16, InceptionV3, and ResNet18 as benchmark models, given their proven efficacy and widespread application in image classification tasks^31^. These well-established deep learning architectures were chosen to provide a robust baseline, facilitating a thorough evaluation of our proposed model’s performance in classifying pathological images.

## Results

We developed and evaluated an end-to-end iMDPath for cancer analysis and grade prediction based on pathological images. This model is specifically designed to address critical challenges, including limited data availability, annotation variation, and the need for model interpretability in clinical applications.

### Generative Model Data Augmentation by iMDPath-Aug

To mitigate the issues of limited data and inconsistent annotations in cancer pathological images, we optimized the generative models for data augmentation in the iMDPath-Aug module, with a particular focus on the VQ-VAE. The introduction of discrete encoding in the VQ-VAE model significantly enhanced the data compression and reconstruction process, particularly for pathological images with discrete structural features. This approach was found to outperform other models in terms of representational capacity for handling complex pathological image structures. We first assessed the impact of the VQ-VAE model on data augmentation by comparing its performance against traditional data preprocessing techniques and the VAE on the BOT and SEED datasets. The results from training with the VQ-VAE model demonstrated notable advantages in both model convergence and classification accuracy.

The VQ-VAE model exhibited excellent convergence, as indicated by the loss curve (**Figures 2A, 3A, 4A, 5A**). This stable convergence curve indicates that the VQ-VAE model achieved efficient data processing without overfitting, demonstrating strong generalization capability. Both training and validation losses plateaued at approximately 0.01, indicating high fidelity in data reconstruction and minimal quantization error. Following data augmentation with the VQ-VAE model, the classification performance improved significantly (**Table 1**). The Swin-B model, enhanced by the VQ-VAE-augmented images, achieved remarkable results on the BOT dataset, with an accuracy of 0.957 and a maximum AUC of 0.995. Similarly, on the SEED dataset, the Swin-B model, optimized with VQ-VAE data augmentation, achieved an accuracy of 0.980 and an AUC of 0.999. These results highlight the effectiveness of the VQ-VAE model in enhancing both classification accuracy and model stability.

**Table 1.**
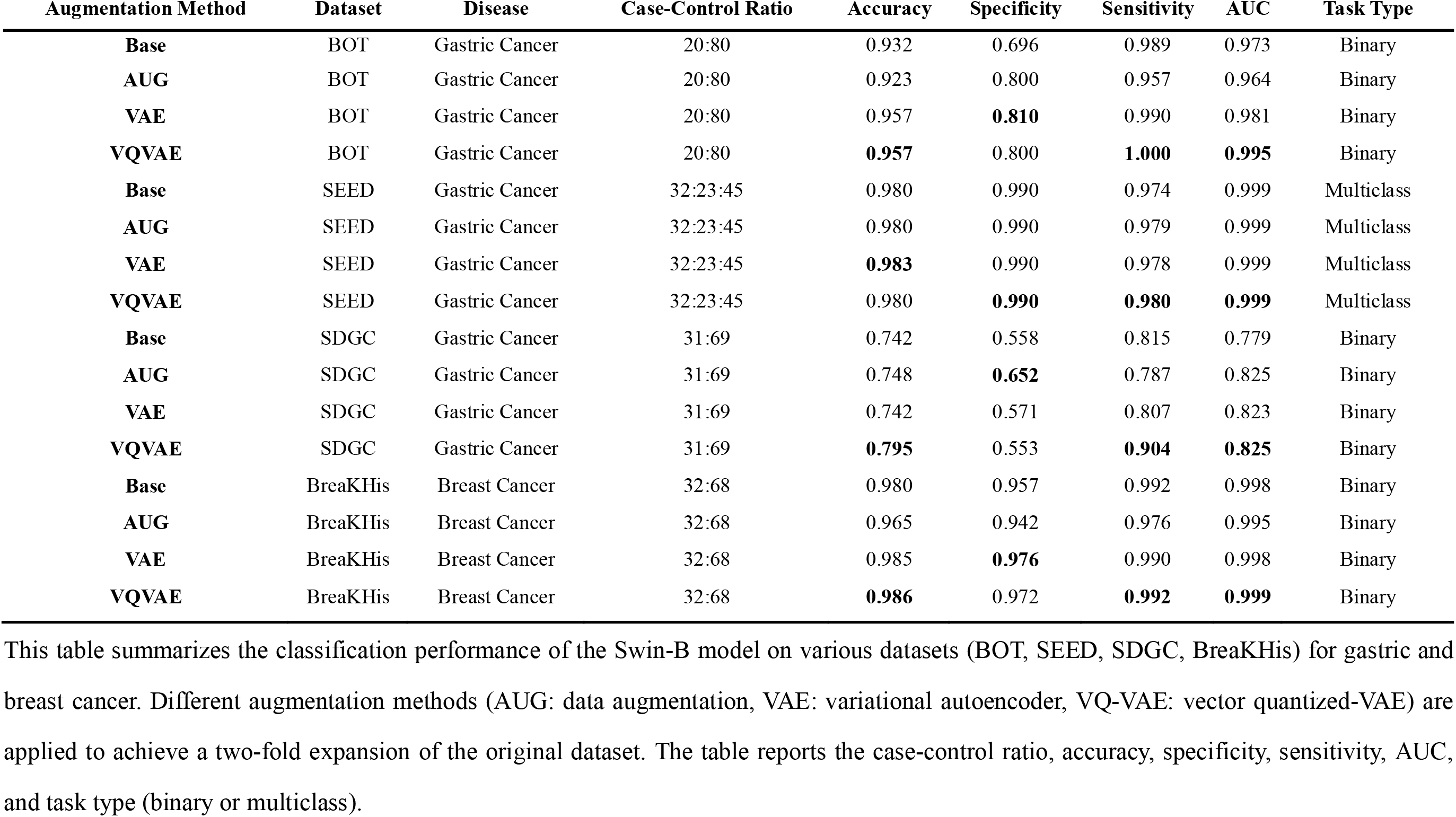
Performance of different augmentation techniques with the Swin-B model in iMDPath framework across different datasets.

**Fig. 2.**
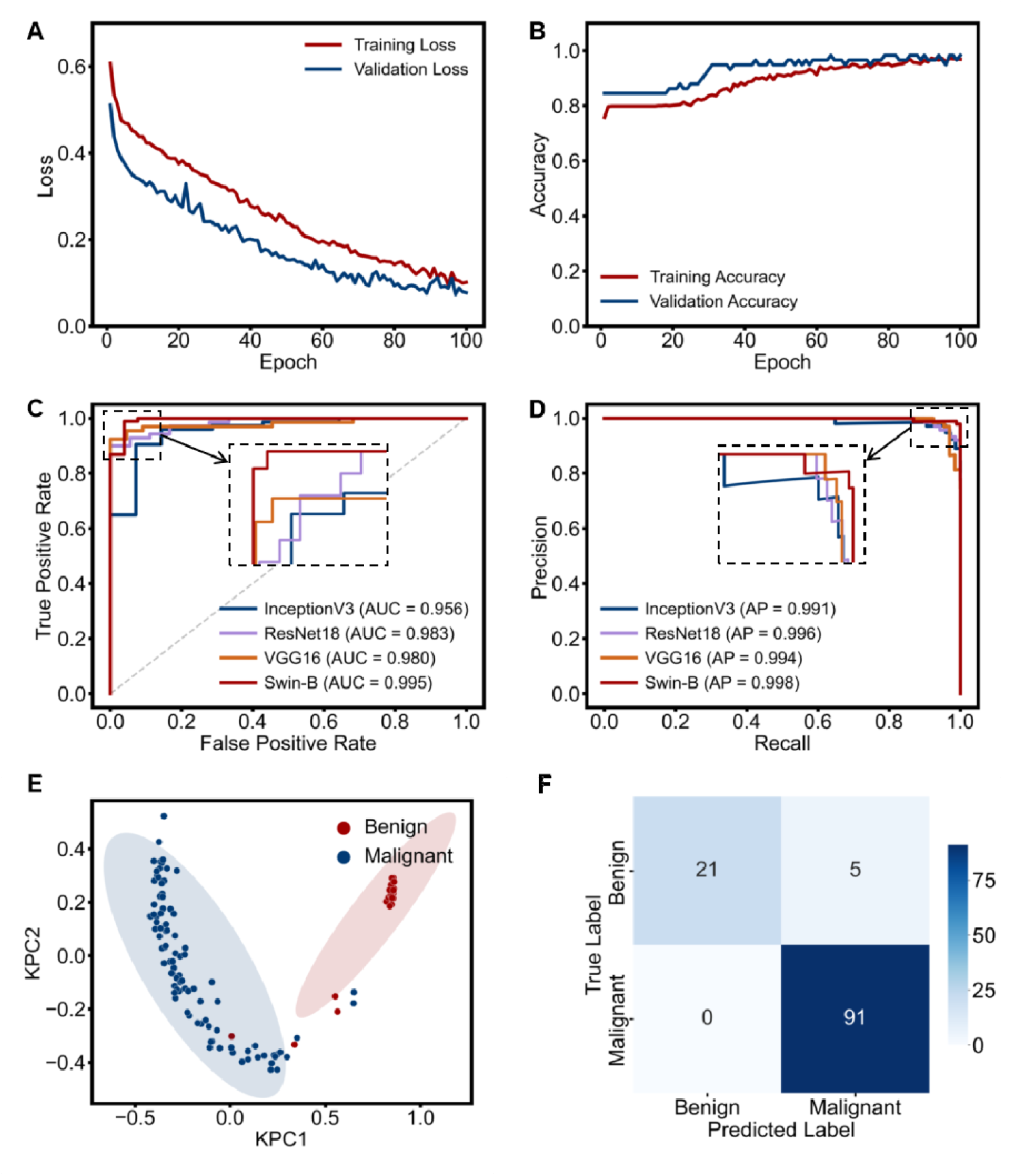
The convergence and classification performance of iMDPath in the BOT Dataset. Panel A shows loss curves showing the model’s performance along the training and validation process, reflecting consistent convergence. Panel B shows the accuracy curves illustrating the improvement in classification performance over epochs. Panel C shows AUC curves indicating the area under the curve for model efficacy across multiple models. Panel D shows the precision-recall curves depicting the balance between precision and recall for various models. Panel E shows the latent embeddings of the iMDPath-Pred module for predicting the outcome. Panel F shows the confusion matrix for model predictions, demonstrating strong classification outcomes.

To validate the performance of the VQ-VAE model on a more diverse set of gastric cancer images, we applied the VQ-VAE data augmentation to the SDGC dataset. The Swin-B model, optimized by the VQ-VAE-augmented images, achieved an accuracy of 0.795 and an AUC value of 0.825. Although the performance was slightly lower compared to the BOT and SEED datasets, these results still underscore the potential of the VQ-VAE model in real-world clinical applications, where image quality and annotation consistency can vary. To evaluate the generalization capability of the VQ-VAE model, we applied it to the BreaKHis dataset consisting of breast cancer images. The loss curves of the VQ-VAE model showed consistent behaviour, stabilizing over time and indicating the model’s robustness in preventing overfitting. The Swin-B model, optimized by the VQ-VAE-enhanced images, achieved 0.986 accuracy and an AUC value of 0.999. The significant improvement in classification performance on this dataset further demonstrates the VQ-VAE model’s excellent generalization capabilities across different cancer types.

### Prediction Performance by iMDPath-Pred

When evaluating the classification performance of various models on pathological images, we observed that the Swin-B model in the iMDPath-Pred module consistently outperformed convolutional models such as VGG16, ResNet18, and InceptionV3 in terms of classification accuracy (**Table 2**). The Swin-B model improves computational efficiency and enhances classification performance by leveraging a sliding window mechanism, which combines local perception with global dependencies.

**Table 2.** Performance comparison of Swin-B in iMDPath framework and other models with VQ-VAE enhancement on pathological datasets.

| Model | Dataset | Disease | Case-Control Ratio | Accuracy | Specificity | Sensitivity | AUC | Task Type |
| --- | --- | --- | --- | --- | --- | --- | --- | --- |
| **VGG16** | BOT | Gastric Cancer | 20:80 | 0.943 | **0.909** | 0.955 | 0.980 | Binary |
| **ResNet18** | BOT | Gastric Cancer | 20:80 | 0.932 | 0.722 | 0.986 | 0.983 | Binary |
| **InceptionV3** | BOT | Gastric Cancer | 20:80 | 0.932 | 0.714 | 0.973 | 0.956 | Binary |
| **Swin-B** | BOT | Gastric Cancer | 20:80 | **0.957** | 0.800 | **1.000** | **0.995** | Binary |
| **VGG16** | SEED | Gastric Cancer | 32:23:45 | 0.952 | 0.977 | 0.958 | 0.997 | Multiclass |
| **ResNet18** | SEED | Gastric Cancer | 32:23:45 | 0.969 | 0.985 | 0.970 | 0.998 | Multiclass |
| **InceptionV3** | SEED | Gastric Cancer | 32:23:45 | 0.969 | 0.986 | 0.960 | 0.997 | Multiclass |
| **Swin-B** | SEED | Gastric Cancer | 32:23:45 | **0.980** | **0.990** | **0.980** | **0.999** | Multiclass |
| **VGG16** | SDGC | Gastric Cancer | 31:69 | 0.775 | 0.310 | **0.955** | 0.823 | Binary |
| **ResNet18** | SDGC | Gastric Cancer | 31:69 | 0.770 | 0.486 | 0.897 | 0.778 | Binary |
| **InceptionV3** | SDGC | Gastric Cancer | 31:69 | 0.795 | **0.568** | 0.888 | 0.788 | Binary |
| **Swin-B** | SDGC | Gastric Cancer | 31:69 | **0.795** | 0.553 | 0.904 | **0.825** | Binary |
| **VGG16** | BreaKHis | Breast Cancer | 32:68 | 0.974 | **0.980** | 0.962 | 0.997 | Binary |
| **ResNet18** | BreaKHis | Breast Cancer | 32:68 | 0.962 | 0.932 | 0.976 | 0.992 | Binary |
| **InceptionV3** | BreaKHis | Breast Cancer | 32:68 | 0.962 | 0.940 | 0.973 | 0.993 | Binary |
| **Swin-B** | BreaKHis | Breast Cancer | 32:68 | **0.986** | 0.972 | **0.992** | **0.999** | Binary |

This table summarizes the classification performance of the VQ-VAE-enhanced models (VGG16, ResNet18, InceptionV3, Swin-B) on various datasets (BOT, SEED, SDGC, BreaKHis) for gastric and breast cancer. The table reports the case-control ratio, accuracy, specificity, sensitivity, AUC, and task type (binary or multiclass).

During training, the Swin-B model exhibited excellent convergence trends, as indicated by the loss curve and the accuracy curve (**Figures 2A, 2B**). In terms of classification performance, the Swin-B model, when optimized with VQ-VAE-augmented data, achieved remarkable results. On the BOT dataset, the Swin-B model achieved optimal performance with peak values of 0.957 for accuracy, 0.995 for AUC, and 0.998 for AP (**Figures 2C, 2D**). To further validate the model’s discriminative power, we analyzed the latent embeddings of the iMDPath-Pred model, which revealed distinct clusters for different disease categories, demonstrating the model’s ability to capture critical pathological features (**Figure 2E**). The confusion matrix for this model revealed high precision and recall across all categories and correct classifications substantially outnumbered misclassifications. This indicates strong discriminative capabilities (**Figure 2F**). The model achieved 80.8% sensitivity for benign cases, with 100.0% specificity for malignant samples.

Similarly, when applied to the SEED dataset, the Swin-B model exhibited excellent convergence (**Figure 3A**), with the loss curve significantly decreasing within the first 20 epochs and then gradually stabilizing. The training and validation loss curves showed consistent trends, confirming the model’s robust generalization capability. As shown in **Figure 3B**, the accuracy on both training and validation sets continued to improve, achieving a stable high accuracy after several iterations, indicating superior feature extraction and classification abilities. When enhanced by the VQ-VAE model, the Swin-B model demonstrated superior performance with an accuracy of 0.980, AP of 0.998, and AUC of 0.999, surpassing the convolutional models by 1.1%-2.8% in accuracy (**Figures 3C, 3D**). The latent embeddings analysis for the SEED dataset (**Figure 3E**) revealed distinct clusters for normal, tubular, and mucinous conditions. The confusion matrix revealed high classification accuracy, with significantly more correctly classified samples than misclassifications, further supporting the model’s robust performance (**Figure 3F**). Multiclass accuracy exceeded 0.976 across all subtypes, with minimal misclassifications: three tubular samples misclassified as mucinous and four mucinous samples misclassified as tubular, suggesting strong overall performance but slight challenges in distinguishing these closely related categories.

**Fig. 3.**
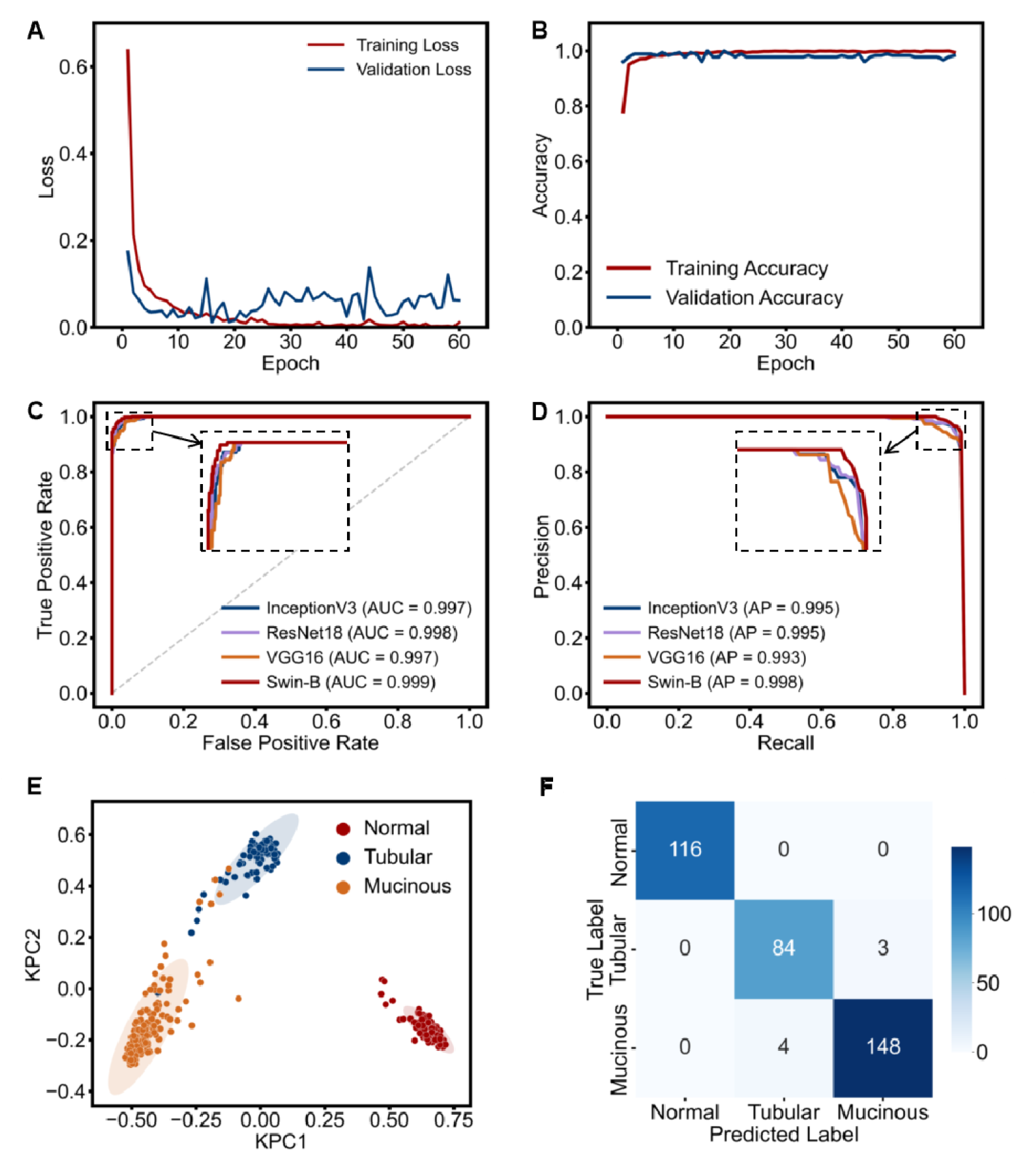
The convergence and classification performance of iMDPath in the SEED dataset. Panel A shows the loss curves that represent the stability of the model’s training and validation performance. Panel B shows the accuracy curves that indicate improvements in classification performance over time. Panel C shows the ROC curves reflecting robust diagnostic performance, with high AUC values. Panel D shows the precision-recall curves demonstrating strong classification ability across different thresholds. Panel E shows the latent embeddings of the iMDPath-Pred module for predicting the outcome. Panel F shows the confusion matrix confirming accurate classification of benign and malignant cases.

To evaluate the performance of the Swin-B model in a real-world clinical setting, we tested it on the SDGC dataset. The loss curve for the Swin-B model demonstrated consistent convergence (**Figure 4A**), and the accuracy curve showed a higher convergence speed and improved final accuracy, confirming the effectiveness of the model architecture and optimization techniques (**Figure 4B**). Upon integration with VQ-VAE, this architecture achieved superior performance metrics, with an accuracy of 0.795, AP of 0.908, and maximum AUC of 0.825 (**Figures 4C, 4D**). The latent embeddings analysis for the SDGC dataset (**Figure 4E**) revealed distinct clusters for benign and malignant conditions. The confusion matrix revealed fewer misclassifications, especially in key categories, validating the model’s capability to handle real-world data with less annotation consistency (**Figure 4F**). The model achieved 79.7% sensitivity for malignant cases, with 55.4% specificity for benign samples. The relatively low number of misclassifications demonstrates the model’s robustness in distinguishing between the two categories.

**Fig. 4.**
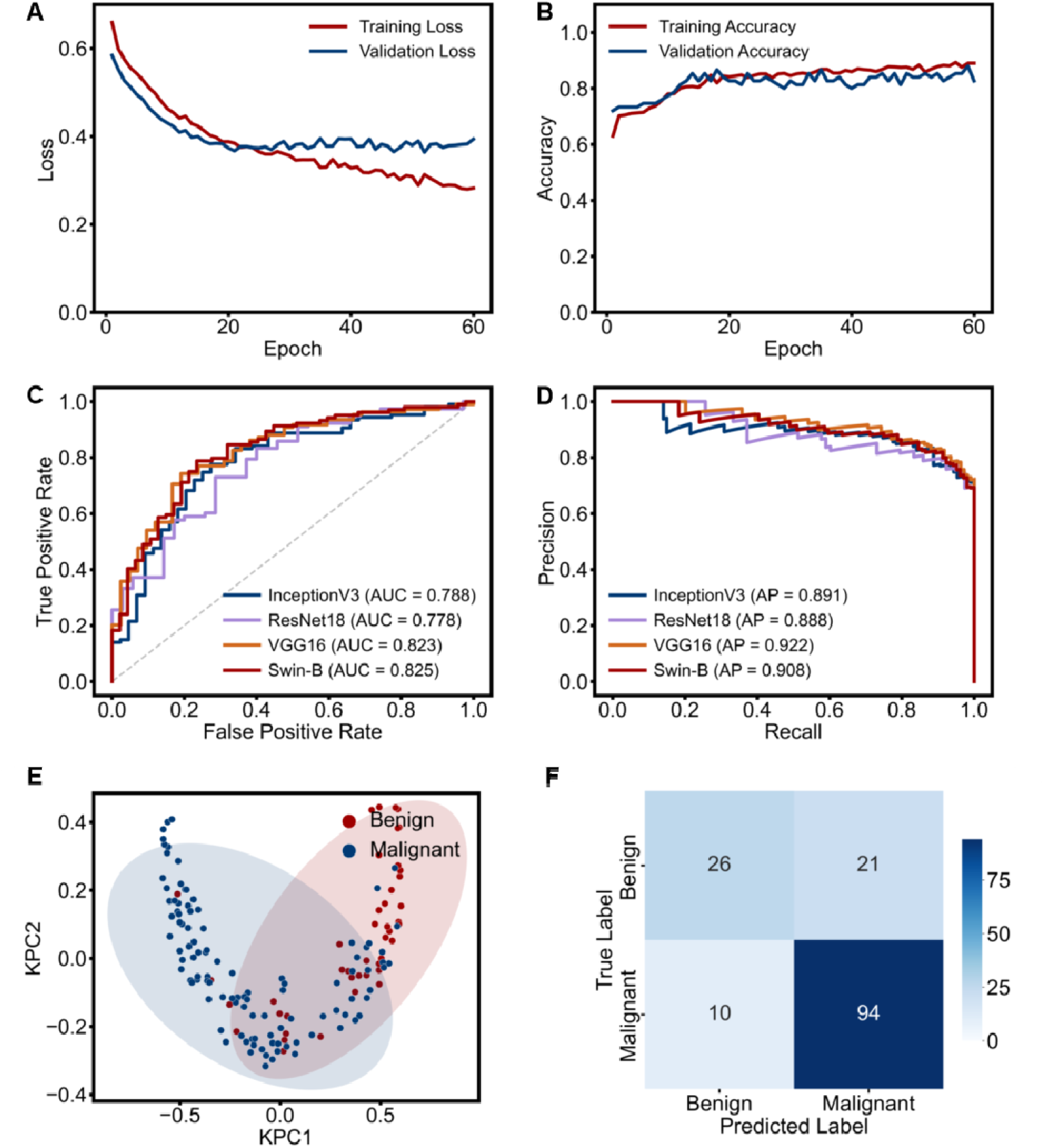
The convergence and classification performance of iMDPath in the SDGC dataset. Panel A shows the loss curves that represent the stability of the model’s training and validation performance. Panel B shows the accuracy curves that indicate improvements in classification performance over time. Panel C shows the ROC curves reflecting robust diagnostic performance, with high AUC values. Panel D shows the precision-recall curves demonstrating strong classification ability across different thresholds. Panel E shows the latent embeddings of the iMDPath-Pred module for predicting the outcome. Panel F shows the confusion matrix confirming accurate classification of benign and malignant cases.

Finally, to assess the generalization capability of the Swin-B model across various cancer types, we tested it on the BreaKHis dataset of breast cancer tissue images. The model’s loss and accuracy curves demonstrated excellent convergence and stable high accuracy (**Figures 5A, 5B**), highlighting the effectiveness of the Swin-B architecture and the importance of data preprocessing and augmentation strategies in optimizing model performance. Notably, after incorporating VQ-VAE data augmentation, the model achieved remarkable results with maximum performance indicators: 0.986 accuracy, AUC of 0.999, and AP of 1.000 (**Figures 5C, 5D**). The latent embeddings for the BreaKHis dataset (**Figure 5E**) showed well-defined clusters, underlining the model’s ability to learn clinically meaningful representations. The confusion matrix for this dataset showed significant classification accuracy across all sample types, with a substantially higher number of correctly classified samples compared to misclassifications, indicating the model’s reliability and strong performance in complex classification tasks (**Figure 5F**). The model achieved 0.972 sensitivity (494 cases) and 99.2% specificity (1065 cases) and very few benign fibroadenomas misclassified as malignant (9 cases).

**Fig. 5.**
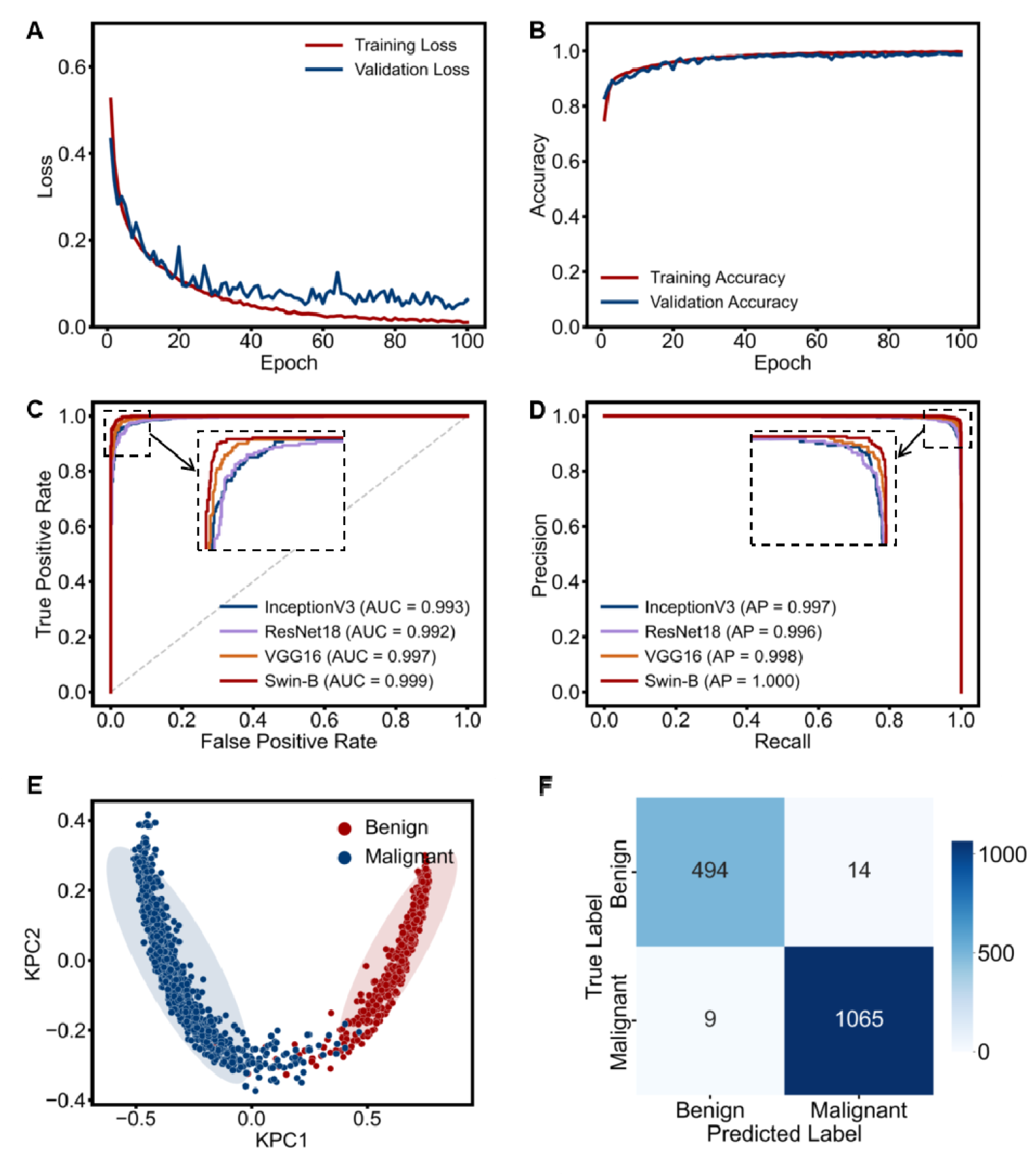
The convergence and classification performance of iMDPath in BreaKHis dataset. Panel A shows the loss curves that represent the stability of the model’s training and validation performance. Panel B shows the accuracy curves that indicate improvements in classification performance over time. Panel C shows the ROC curves reflecting robust diagnostic performance, with high AUC values. Panel D shows the precision-recall curves demonstrating strong classification ability across different thresholds. Panel E shows the latent embeddings of the iMDPath-Pred module for predicting the outcome. Panel F shows the confusion matrix confirming accurate classification of benign and malignant cases.

### Visualizing the Latent Embedding Enhances Interpretability by iMDPath-Vis

The iMDPath-Vis module provides the heatmap visualizations to the parameter weights across the pathological regions. **Figures 6-8** show the example images among multiple disease classes to illustrate the enhanced interpretability for the iMDPath framework. The model effectively identifies and highlights key lesion areas associated with cancer, even within complex tissue structures.

**Fig. 6.**
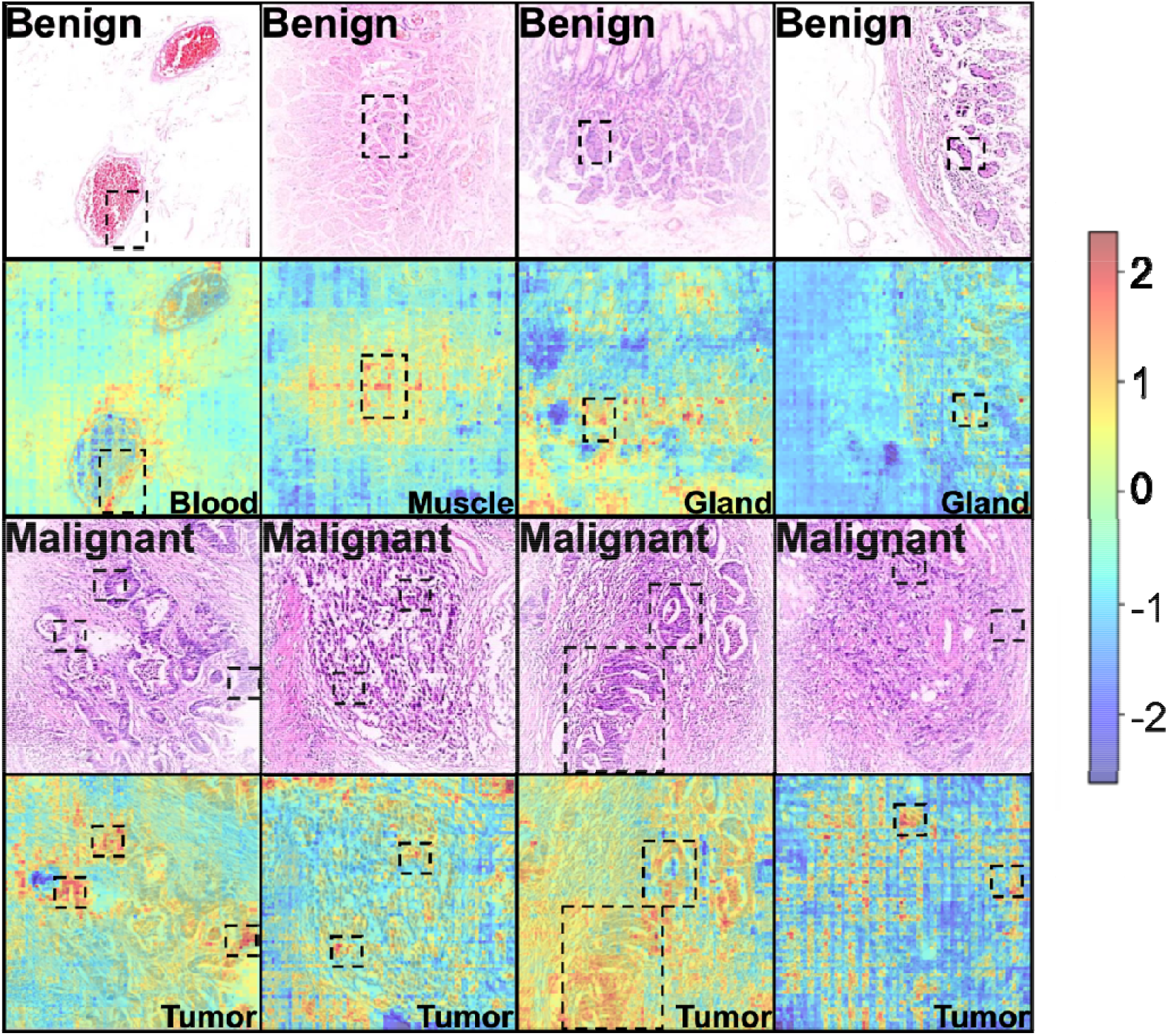
The latent embeddings and interpretable visualization for pathological images in the BOT dataset. The heatmaps illustrate attention regions in benign and malignant cases, with FullGrad-based visualization highlighting key tissue regions in the pathological images.

Attention heatmaps are visualized through a color gradient, where red indicates high attention weights and blue represents low relevance. In **Figure 6 and Supplementary Figure**, benign gastric cases show heatmaps tracing intact vascular networks and glandular structures. Malignant cases highlight cancer cell clusters at tumor-stroma interfaces, aligning with the vascular-glandular niche disruption concept^32^. **Figure 7** demonstrates subtype-specific attention patterns in mucinous gastric tumors: heatmaps localize to fragmented mucin pools with disrupted epithelial continuity, indicative of mucin-barrier collapse^33^. Cross-cancer validation in **Figure 8** shows benign breast lesions draw attention to luminal structures and collagen fibers. Malignant cases prioritize disordered cancer nests with nuclear atypia, consistent with WHO diagnostic standards.

**Fig. 7.**
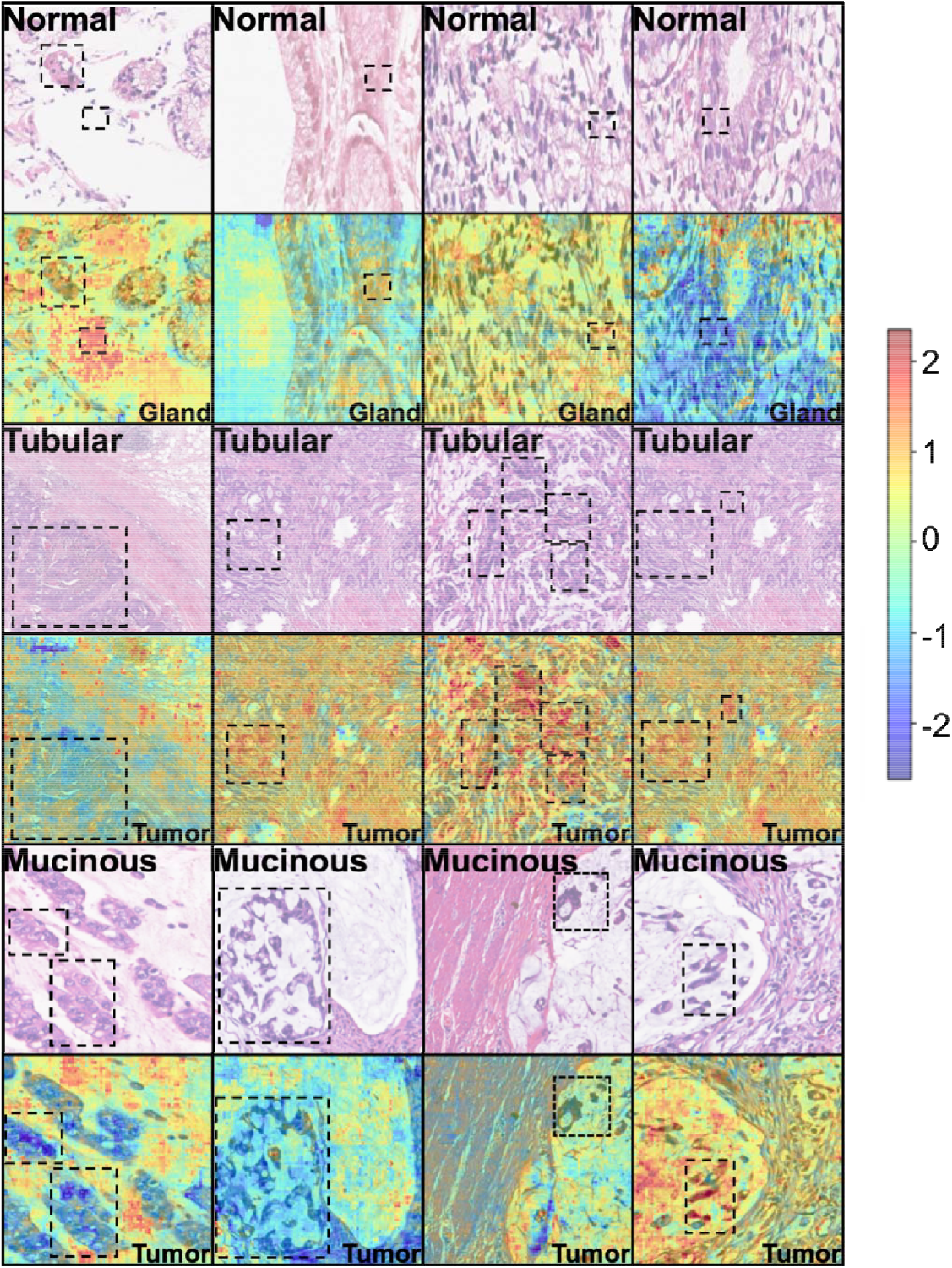
The latent embeddings and interpretable visualization for pathological images in the SEED dataset. Example pathological images with attention heatmaps focusing on diagnostically relevant areas. The visualization effectively isolates tumor-rich regions in mucinous and tubular samples.

**Fig. 8.**
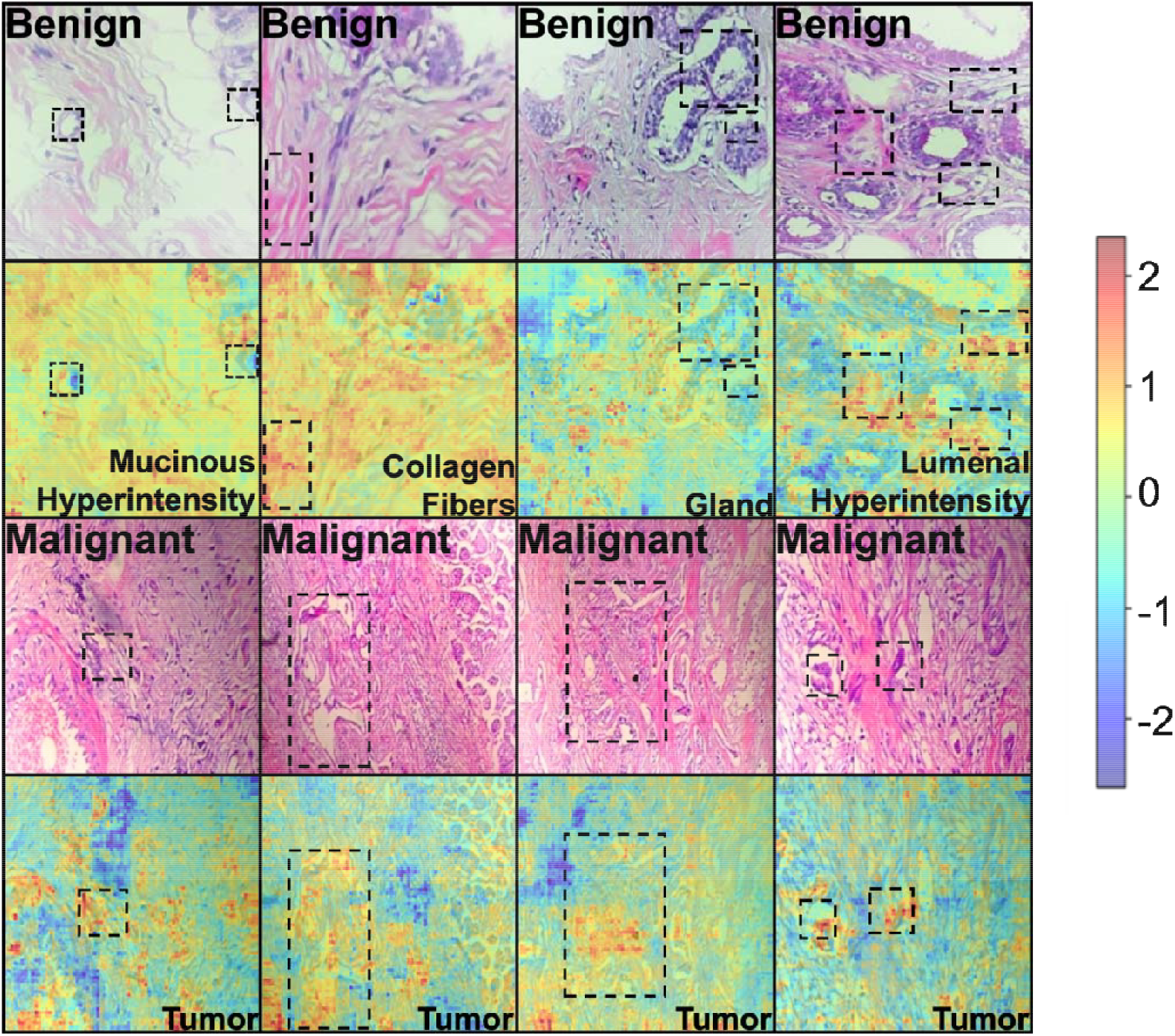
The latent embeddings and interpretable visualization for pathological images in the BreaKHis dataset. The heatmaps highlight critical regions associated with cancerous tissue, enhancing interpretability by emphasizing diagnostically significant areas in breast cancer samples.

By visually emphasizing these critical areas, the model provides valuable insights into the underlying pathological features and decision-making processes, facilitating a better understanding of the model’s feature selection across different cancer types. Overall, the visualization approach presented in this study not only achieves remarkable accuracy in classifying pathological images of various cancers but also holds significant potential for clinical applications.

## Discussion

This study presents iMDPath, a novel end-to-end framework uniquely integrating image data augmentation, disease grade prediction, and interpretability for efficient and accurate pathological image analysis. The synergistic interaction of its modules, iMDPath-Aug, iMDPath-Pred, and iMDPath-Vis, addresses critical challenges in digital pathology, paving the way for enhanced clinical utility of deep learning models.

The VQ-VAE generative model within the iMDPath-Aug effectively expands and enhances the dataset sample sizes, improving disease outcome prediction by compressing and reconstructing pathological images through a discrete encoding mechanism. This significantly bolsters the generalization capability of the iMDPath framework, particularly evident in the SDGC and BreaKHis datasets. Specifically, the introduction of VQ-VAE yielded substantial improvements in prediction accuracy (5.3% increase in the SDGC dataset), highlighting the importance of generative models in pathological image data processing and their potential to enhance model performance, especially when dealing with limited and heterogeneous datasets.

Compared to state-of-the-art convolutional neural networks, including VGG16, ResNet18, and InceptionV3, the iMDPath-Pred module demonstrates superior accuracy and stability in pathological image classification tasks. The Swin-B architecture, leveraging a sliding window mechanism, integrates local perception with global dependencies, reducing computational complexity and enabling efficient processing of high-resolution pathological images. Notably, after data augmentation by the iMDPath-Aug module, iMDPath achieved remarkable performance improvements across multiple datasets. For instance, the model reached an accuracy of 0.989 and an AUC value approaching 1.000 in the SEED and BOT datasets, showcasing its strong discriminatory power and wide applicability. Importantly, these results were consistently observed across independent test sets, further validating the robustness of the framework. These results show that iMDPath achieves better performance on different datasets, such as the average AUC improvement is 2.2%, 0.2%, 2.9%, 0.5% higher than VGG16, ResNet18, and InceptionV3 on the BOT, SEED, SDGC and BreaKHis datasets, respectively.

Enhanced by the FullGrad and occlusion sensitivity analysis, the iMDPath-Vis module provides multi-level interpretability by identifying regions of interest in the input images and quantifying the contribution of each pixel to the final prediction. This level of interpretability is crucial for clinical decision-making, providing pathologists with intuitive image interpretations and insights into the model’s feature selection process. For example, the visualization consistently highlighted tumor infiltrating lymphocytes (TILs) in gastric cancer samples as key predictive features, aligning with established histopathological knowledge and suggesting potential for iMDPath to aid in identifying prognostically relevant features.

There are a couple of limitations in the study. While the iMDPath-Aug module addresses data insufficiency, the quality of the generated images may suffer from some noise, which could influence model stability, especially when the dataset sizes are very limited and the variations of pathological images are higher. To mitigate this, we implemented rigorous quality control measures during data augmentation and employed techniques such as adversarial training to improve the realism of the generated images. Future work will focus on exploring alternative generative models and incorporating domain-specific knowledge to further enhance the quality and diversity of the augmented data. Moreover, the current study primarily focused on gastric and breast cancer datasets. Future studies should evaluate the iMDPath framework on a broader range of cancer types and imaging modalities to assess its generalizability and identify potential adaptations needed for different clinical contexts.

## Conclusions

In conclusion, we have developed iMDPath, a novel end-to-end framework that uniquely integrates data augmentation, accurate prediction, and interpretable visualization for clinical pathological image analysis. iMDPath demonstrates exceptional performance across diverse gastric and breast cancer datasets, and provides pathologists with actionable insights into the model’s decision-making process, highlighting key tissue regions. By simultaneously addressing the challenges of data scarcity and interpretability, iMDPath represents a significant step forward in the application of deep learning to precision oncology. This framework holds promise for enhancing diagnostic accuracy, streamlining pathologist workflows, and ultimately improving patient outcomes through more personalized and effective cancer treatment strategies.

## Data availability

The source code to process data and generate the figures in the manuscript is available on GitHub (https://github.com/mmetalab/iMDPath) and the original datasets are available on Hugging Face (https://huggingface.co/chenqitao).

## Acknowledgment

We thank the high-performance computing service provided by the National Institute of Health Data Science of China, Shandong University.

## Abbreviation

AP: Average precision
AUC: Area under the curve
BOT: Brain of Things
CAM: Class activation map
DL: Deep learning
FN: False negative
FP: False positive
FPR: False positive rate
FullGrad: Full gradient
iMDPath: Interpretable Multi-Task Digital Pathology Model
iMDPath-Aug: Interpretable Multi-Task Digital Pathology Model-Augmentation
iMDPath-Pred: Interpretable Multi-Task Digital Pathology Model-Prediction
iMDPath-Vis: Interpretable Multi-Task Digital Pathology Model-Visualization
LayerNorm: Layer normalization
MLP: Multi-layer perceptron
PR: Precision-recall
ROC: Receiver operating characteristics
ResNet: Residual Neural Network
SGD: Stochastic gradient descent
Swin-B: Swin Transformer-Based
SW-MSA: Shifted window multi-head self-attention
TILs: Tumor infiltrating lymphocytes
TN: True negative
TP: True positive
TPR: True positive rate
VAE: Variational autoencoder
VGG: Visual Geometry Group
ViT: Vision Transformer
VQ-VAE: Vector-quantized variational autoencoder
W-MSA: Window-based multi-head self-attention

