## Supplemental Figure 1 for "iMDPath: Interpretable Multi-task Digital Pathology Model for Clinical Pathological Image Prediction and Interpretation"

Tao Zhang

Cheng Wang

† The authors wish it to be known that, in their opinion, the first two authors should be regarded as joint first authors.
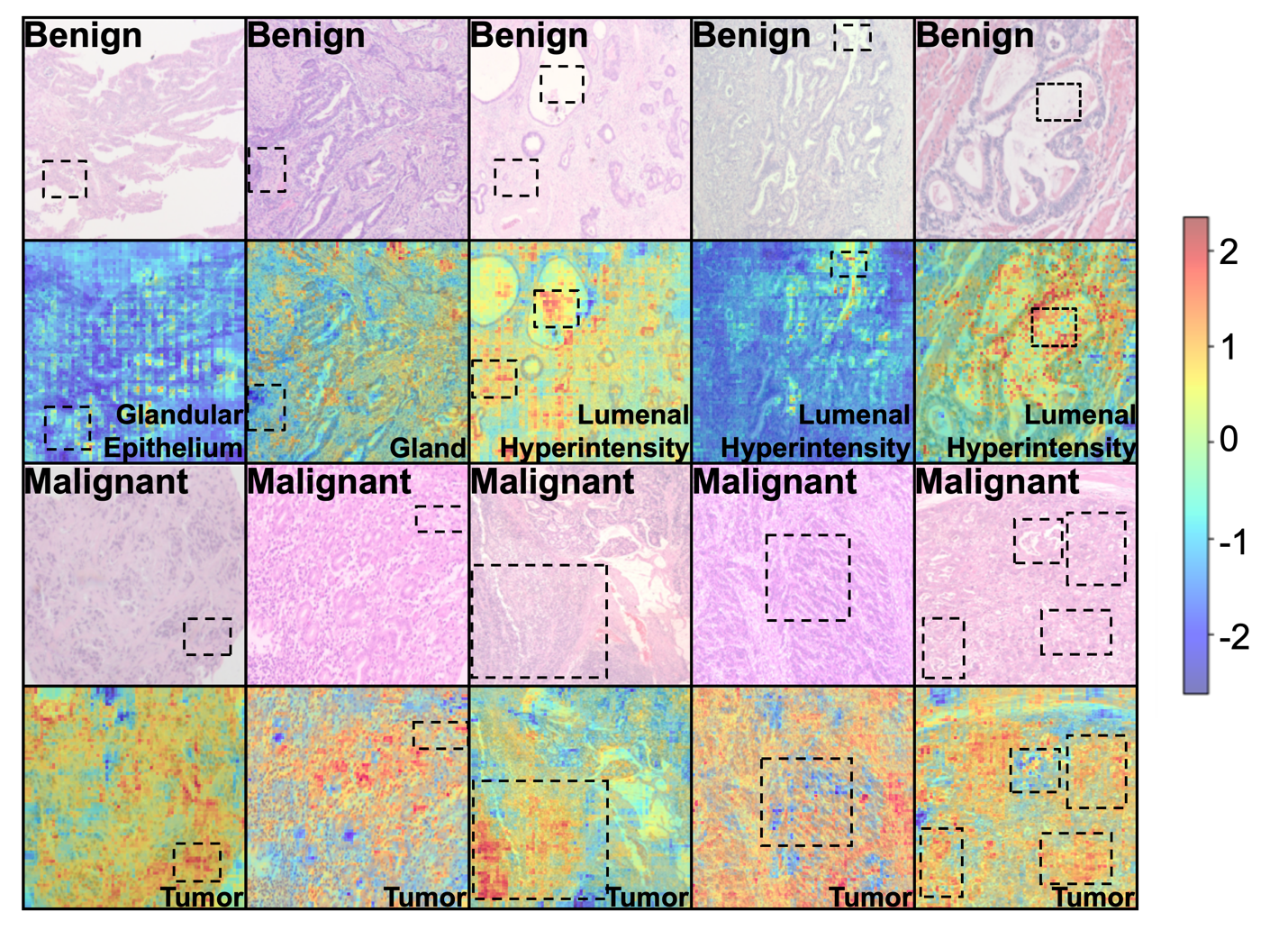

**Figure S1. The latent embeddings and interpretable visualization for pathological images in the SDGC dataset.** The heatmaps highlight critical regions associated with cancerous tissue, enhancing interpretability by emphasizing diagnostically significant areas in gastric cancer samples.
